# [Re] Deep Learning for ECG Analysis: Benchmarks and Insights from PTB-XL

**DOI:** 10.1101/2025.01.27.25321112

**Authors:** Antony M. Gitau, Ayoade Adeyemi, Belal Tavashi, Bjørn-Jostein Singstad

## Abstract

**Reproducibility Summary:** *Scope of Reproducibility:* The authors of the original paper present six benchmark tasks on the previously published PTB-XL dataset, containing, 21837 12-lead ECGs from 18885 patients. They evaluate seven different neural network architectures on the six benchmark tasks. The authors have published all code and claim full reproducibility. In addition, they published code for easy implementation of new models. To validate the claim of reproducibility we implemented a new model and tested it, and the seven models presented by the authors of the original paper, on the six benchmark tasks.

*Methodology:* We used the publicly available code, published by the authors of the original paper, as a starting point for our experiment. Furthermore, we modified the code slightly in order to make it compatible with a cloud-hosted Jupyter Notebook, Google Colab. We ran the experiments using Google Colab Pro, using 32 GB RAM and either 1 x NVIDIA P100 or 1 x NVIDIA T4 GPU.

*Results:* We successfully managed to reproduce the original work and also verified the validity of the main claims of the original paper. In addition, we showed how robust the models were to noise and finally implemented a new model that showed comparable performance with the models proposed in the original paper.

*What was easy:* The publicly available code published by the authors made it easy to reproduce and obtain the same results as reported in their paper.

*What was difficult:* We faced two main issues in this work. (1) running the code in a cloud-hosted jupyter notebook. This was done in order to get access to free or cheap GPUs. (2) Implement own models using the provided template. The description on how to use the base class and the configuration file could have been more detailed.

*Communication with original authors:* Communication with the authors of the original paper was established early in the project and helped us by clarifying some aspects of the work. In the final stage of this project the authors of the original paper were given this manuscript in order to read it and provide feedback.

## 1 Introduction

An electrocardiogram (ECG) is a representation of the electrical system of the heart that can be obtained non-invasively, making it accessible and easy to use. For more than a century, ECG have been used by doctors and cardiologists to diagnose and prognosticate cardiovascular diseases. In the last decade, deep neural networks (DNN) or more specifically; convolutional neural networks (CNN) have shown promising performance in interpreting ECGs. Previous studies have shown that CNNs can be used to detect various diseases from the ECG at cardiologist level performance [1]. Others have shown that CNNs can detect markers in the ECG that are out of scope for a human interpreter, such as detecting paroxysmal atrial fibrillation [2] and determining age and gender from the ECG [3]. However, Strodthoff et al. emphasize that two bottlenecks slow down the development in the field of artificial intelligence and ECG. First, there is a lack of large and open datasets and secondly, the open datasets miss some clearly defined bench-marking tasks with standardized evaluation procedures [4]. To address this, Wagner et al. published an open data set, PTB-XL, containing 21837 ECG records from 18885 patients [5]. Then, Strodthoff et al. proposed six different benchmarking tasks and used seven different state-of-the-art models on the PTB-XL dataset [4].

In this paper, we have replicated the results from Strodthoff et al. who applied seven models on the six different benchmarking tasks [4], by reusing the authors’ open-source Python implementation with some minimal modifications to allow the code to be run in Google Colab. We replicated the results presented in Strodthoff et al. by running the repeated (three times) bootstraps on the test data. We also performed experiments with different levels of noise added to the test ECGs to evaluate the model’s susceptibility and robustness to noise. Finally, we also proposed a new model, Inception Time (TensorFlow implementation). First, we performed a hyperparameter optimization to find the optimal configuration of the model for each of the six benchmark tasks. The models were evaluated according to the code provided in the GitHub repository published by Strodthoff et al.

## 2 Scope of reproducibility

Strodthoff et al. propose six benchmarking tasks for ECG classification using the PTB-XL dataset. Furthermore, they applied seven different state-of-the-art deep learning-based time series classification algorithms on these benchmark tasks and presented the result in terms of area under the receiver operating characteristic (AUROC) curve. Based on this work the authors have formulated four main claims:

1. Reproducible results by providing the full source code.
2. A framework for easy implementation of new model architectures.
3. Providing a reliable assessment of transfer learning in the ECG context and demonstrating the promising prospects of transfer learning from PTB-XL to other ECG classification datasets in the small dataset regime.
4. Providing evidence for the phenomenon of hidden stratification, a first evaluation of the diagnosis likelihood information provided within the dataset in comparison to model uncertainty and presenting an outlook to possible applications of interpretability methods in the field.

In this replication paper, we have chosen to focus on the two first claims. To test the first claim, we will run the code multiple times and compare it with the published results in Strodthoff et al. to test reproducibility. To test the second claim, we will implement a model that has shown promising performance in similar ECG classification tasks to assess the ease of implementing a new model using the proposed framework by the original authors. In addition, we conducted a grid search to find the optimal hyperparameters for this model for each of the six tasks. Finally, we also want to evaluate the robustness of the models by adding various levels of noise to the ECGs in the test set prior to prediction and evaluation.

## 3 Methodology

To reproduce the results reported by Strodthoff et al., as well as to implement our proposed model, we started by forking and cloning the open available GitHub repository ^1^. We used the freely available GPUs in Google Colab to train the models, although we had to modify the code slightly to make it work. Particularly, we modified the progress bar module used in the fastai python package to make the code compatible with the Google Colab Notebook ^2^.

### 3.1 Model descriptions

The following seven models were implemented and tested on the benchmark tasks in Strodthoff et al., and replicated in this paper:

1. A fully convolutional network (fcn_wang) [6]
2. A standard ResNet-based architecture (resnet1d_wang) [6, 7]
3. A ResNet-based architecture inspired by recently improved ResNet architectures such as xResNets (xresnet1d101) [8]
4. Implementation of Inception Time architecture [9] with the use of a concatenation pooling layer
5. Unidirectional LSTM (lstm) [10]
6. Bidirectional LSTM (*lstm_bidir*) [10]
7. A neural network classifier trained on wavelet features (Wavelet+NN) [11]

In addition to the above-mentioned models, the authors of Strodthoff et al. stated that they tested a unidirectional and bidirectional gated recurrent unit (GRU) network. However, the results from the GRU models were not reported in the paper nor on the benchmark leaderboard displayed in the README file of the author’s GitHub repository.

Our Jupyter notebook implementation of the source code downloads the PTB-XL Dataset, the GitHub repository, and from the GitHub repository it imports the necessary Python packages and finally runs the reproduce_results.py file that runs and validates the models as intended by the authors.

We also implemented a new variation of the Inception model [9], which is already implemented by Strodthoff et al., but in contrast to Strodthoff et al.’s implementation, which was based on PyTorch, we here used TensorFlow. In addition, we optimized the hyperparameters of the model for each specific benchmark task. The hyperparameter tuning process is explained in depth later in a subsection of this chapter.

The source code for our Jupyter Notebook implementation is openly available on GitHub ^3^.

### 3.2 Datasets

The dataset used in this study was the PTB-XL dataset presented by Wagner et al [5]. The dataset is stored on PhysioNet [12, 13], a data bank for physiological signals ^4^. The dataset contains 21837 ECG recordings from 18885 patients and also comes with a large variety of machine- and cardiologist-annotated labels and diagnoses which in Strodthoff et al. were used to propose six different benchmark tasks.

Benchmark tasks — The following benchmark tasks were proposed by Strodthoff et al.:

- *diagnostic*
- *superdiagnostic*
- *sub-diagnostic*
- *form*
- *rhythm*
- *all*

The benchmark task named *diagnostic* refers to classifying all the available diagnostic statements (40) in the dataset. *Superdiagnostic* refers to the five main classes in the data set; Normal ECG (NORM), Conduction Disturbance (CD), Myocardial Infarction (MI), Hypertrophy (HYP), ST/T change (STTC), while *sub-diagnostic* consider the 23 subclasses based on the 5 *Superdiagnostic* classes. The *form*-benchmark contains 19 classes describing the morphology of the ECG, such as abnormal QRS, inverted T-waves, etc. The *rhythm*-benchmark contains 12 classes and describes the ECG rhythms, such as sinus rhythm, sinus bradycardia, atrial flutter, etc. Finally, the benchmark task called *all* refers to the union of all diagnostic, rhythm and form statements (70 classes).

#### Data partitioning

As well as publishing the dataset Wagner et al also proposed a pre-defined data partitioning to ensure reproducible results and thereby making the reported results from different models and algorithms more comparable. The dataset was divided into 10 folds, where fold 1-8 should be considered as the training set, fold 9 as the validation set and fold 10 as the test set [5].

#### Preprocessing

The ECG recordings were converted to a WaveForm DataBase (WFDB) format with a resolution of 1 μV/LSB and 500Hz after the acquisition, and for the user’s convenience, due to memory, the ECGs were also downsampled to 100Hz.

In our implementation, we also added a new argument to the function, which adds a desired level of noise to the test data. In this paper, we use this to evaluate how susceptible the models are to noise by monitoring the decline in AUROC.

### Hyperparameters

The configuration and hyperparameters used in the seven models proposed by Strodthoff et al. were kept when we replicated the results in this work. However, the hyperparameters related to the model we propose in this paper, Inception Time, were tuned by doing a grid search on a subset = 10% of the total training set. A separate grid search was performed for each of the six benchmark tasks, resulting in six different sets of model configurations; one for each of the six benchmark tasks. Table 1 shows the parameters and parameter space used in the grid search.

**Table 1.** Hyperparameter search space for the Inception Time model proposed in this paper.

| Parameter | Values |
| --- | --- |
| Epochs | [15,20,25] |
| Batch size | [16, 32, 64] |
| Initial learning rate | [0.001, 0.0001, 0.00001] |
| Learning rate reduction | [yes, no] |
| Model depth | [6, 9, 12] |
| Loss function | [binary cross-entropy, weighted binary cross-entropy] |
| Kernel size | [(20,10,5),(40,20,10),(60,30,15)] |

Figure 1 shows the results of the grid search performed for all six tasks. Parameter values that gave the same score on the test set are stacked horizontally, while higher scores give a higher value on the vertical axis. The configuration that resulted in the highest score on the vertical axis was selected for final training and testing on the test set.

**Figure 1.**
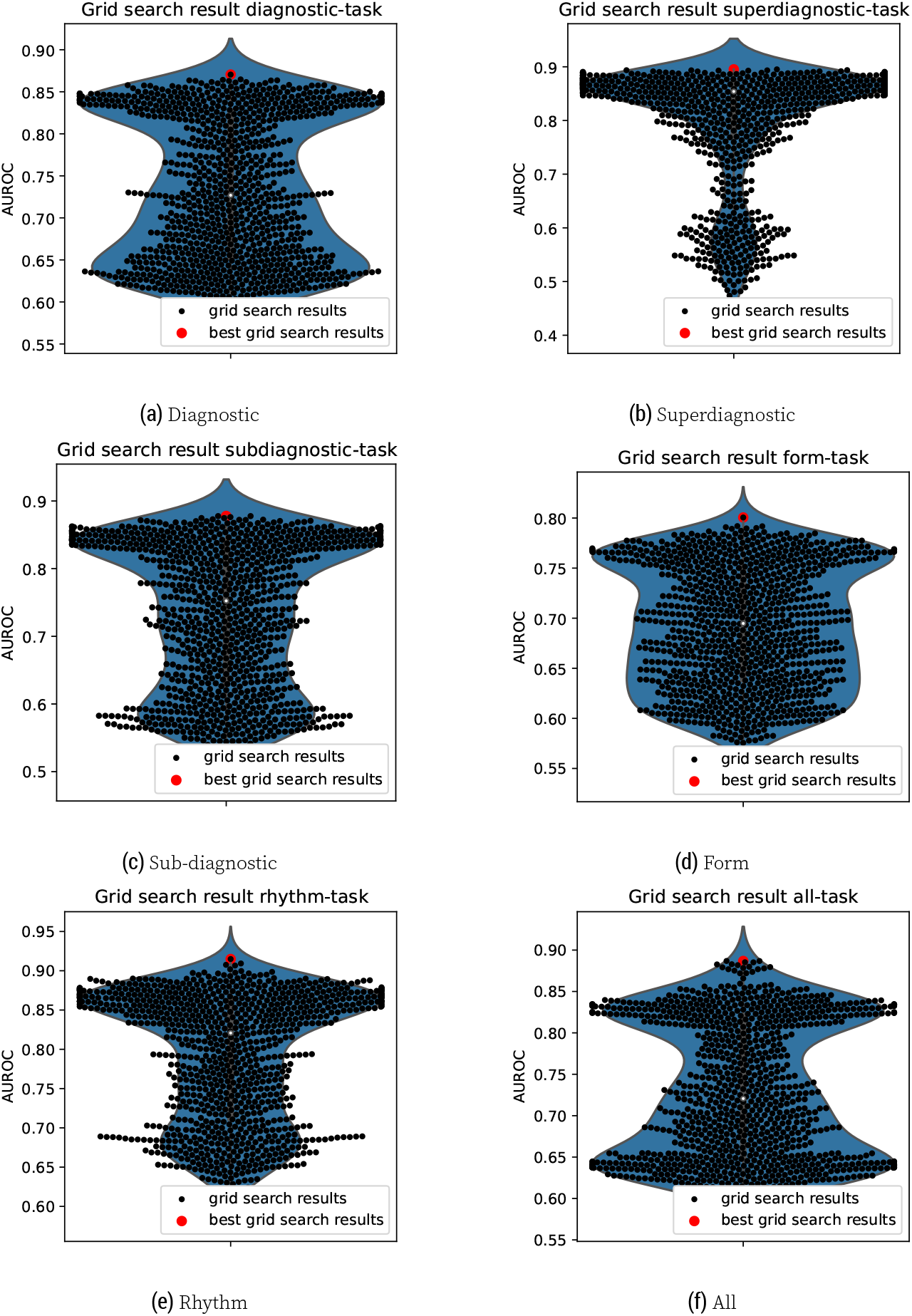
Scores obtained when searching for the optimal model configurations for Inception Time. Each sub-plot represents the scores obtained, using grid search, for one specific benchmark task.

### 3.4 Experimental setup and code

The open available GitHub repository, published by Strodthoff et al, holds a folder named *code*, which includes the Python modules and the code used to run the experiments reported in their work. In addition, the code folder contains templates on how to add new models to make it easier for others to implement new models and test them on the benchmark tasks. Figure 2 shows the files and sub-folders inside the code folder in the GitHub repository. In the configs folder, inside the code folder, there are Python files containing specific parameter configurations to use when training the different models. In case someone wants to implement a new model and apply it to the benchmark tasks they should specify configurations of the new model in the file named your_configs.py.

**Figure 2.**
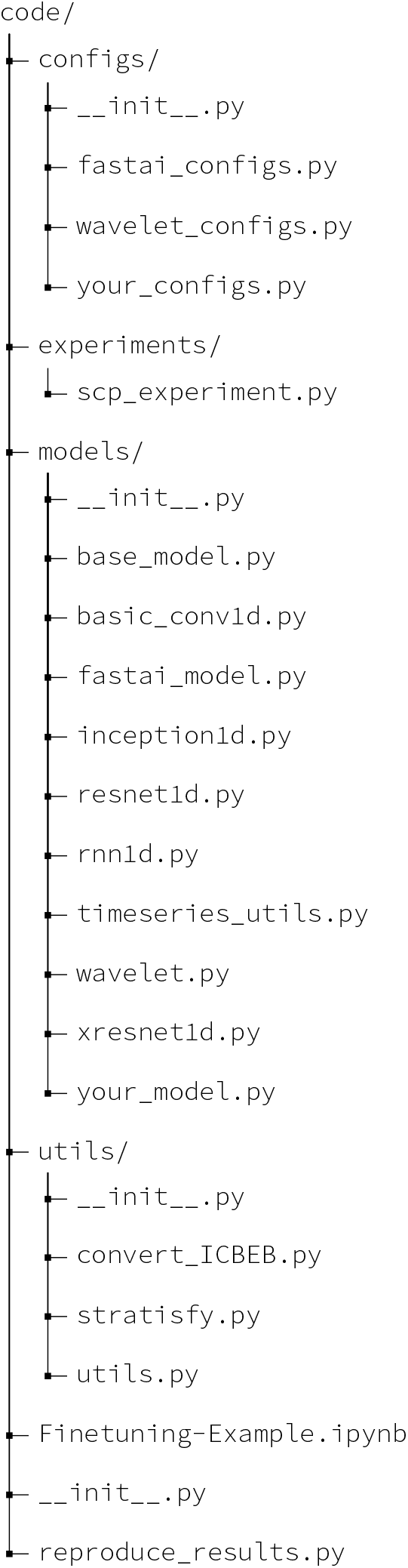
The folder structure within the code folder in GitHub repository published by Strodthoff et al. [4]

Furthermore, in the experiment folder, there is one file named `scp_experiment.py` which specifies the procedure of how the dataset is loaded into the model during training and testing and how the predictions are being stored and evaluated. If someone wants to propose a new model, they should specify the model name, defined in the configuration file, and import the module that includes the proposed model and assign the model to a variable in the `perfom()` function. The utils folder contains four files. `init .py` is necessary to define the util folder as a module, but do not contain any information. The `convert_ICBEB.py` file is used to convert the data from a second database, China Physiological Signal Challenge 2018, to the same format as PTB-XL. `stratisfy.py` is the file that was used by Strodthoff et al to partition the data into 10 folds. `utils.py` contains Python modules used by the other Python files in the folder. `Finetuning-Example.ipynb` is a Jupyter notebook that shows an example of how one can fine-tune new models and validate performance before submitting models to be validated on the benchmark tasks. Finally, the file named `reproduce\_results.py` specifies how to run all selected models through the selected benchmark task.

To reproduce the results from Strodthoff et al., the default settings in the code folder were used, while some modifications to the code had to be done to implement our proposed model. The results and the ranking of the models are based on the area under the receiver operating curve (AUROC). An AUROC score is reported for each model on each benchmark task.

### 3.5 Computational requirements

To run the experiments it is recommended to use GPUs. We did not have access to physical GPUs, so to get access to GPUs, we used Google Colab. To run Python code in Google Colab the code has to be run from a Jupyter notebook. Therefore we had to do some minimal modifications to the original code to make the code run in Google Colab. We used Google Colab with the Pro subscription giving us access to slightly more GPU and RAM compared to the free subscription.

## 4 Results

### 4.1 Results reproducing original paper

The results from running the seven models on the six benchmark tasks are presented in Table 2. In contrast to Strodthoff et al., we here repeated the training and bootstrapping three times to get an even more accurate result. The results are presented as the mean of the three experiments and the background of each value in the table indicates whether the obtained results are inside (green) or outside (red) the 95% confidence interval presented in Strodthoff et al.

**Table 2.**
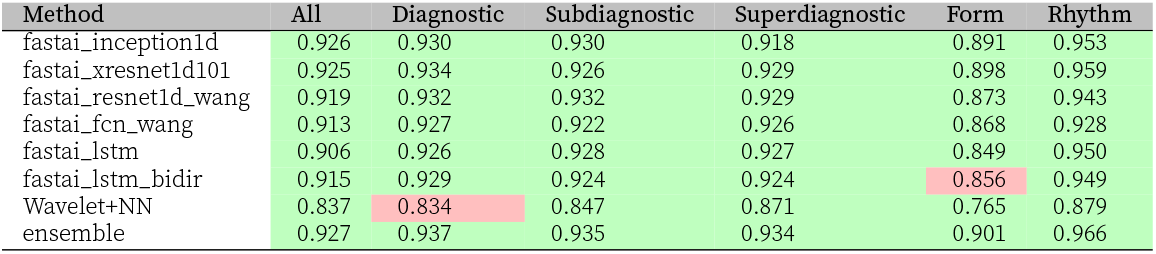
Overall performance, in terms of AUROC, of the ECG classification algorithms proposed by Strodthoff et al. [4] The results are obtained taking the mean of repeated (3 times) bootstrapping on the test set. Table cells with green background indicate that the results obtained are within the confidence interval reported in Strodthoff et al.

### 4.2 Results beyond the original paper

#### Adding noise to ECGs in test data

Figure 4 presents the performance of the seven models in terms of AUROC score on the six different benchmark tasks proposed by Strodthoff et al. when different levels of stochastic noise were added to the test ECG. The stochastic noise was centered around zero with upper and lower boundaries given in terms of the standard deviation, *σ*, of all samples in the ECG test data multiplied by a coefficient *x*. In the experiments shown in Figure 4 *x* were equal to 0, 0.1, 0.5 and 1. Figure 3 shows an example of an ECG with gradually more noise added.

**Figure 3.**
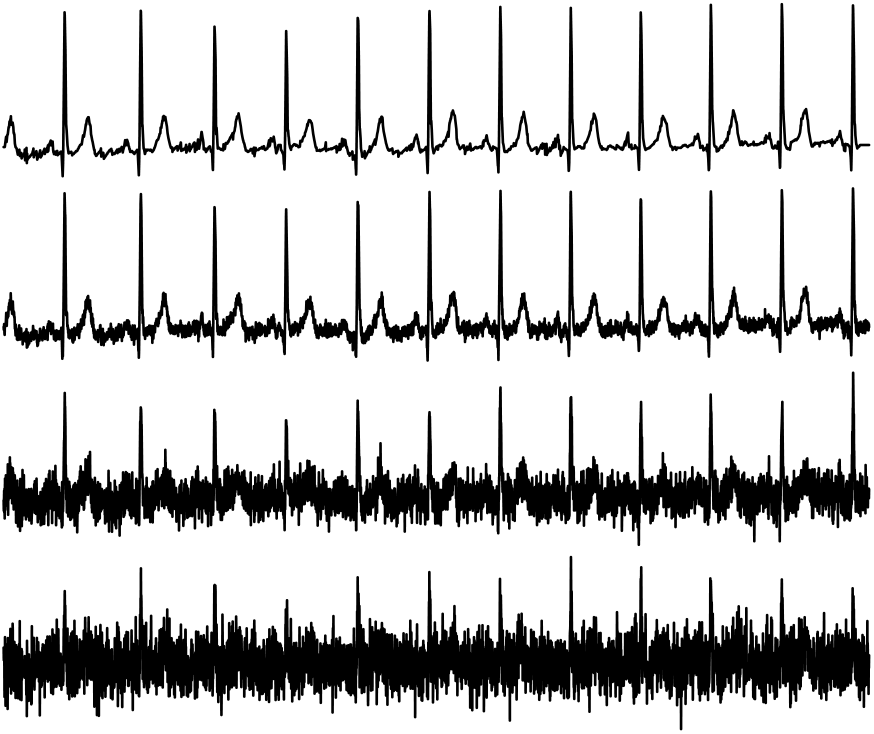
A randomly chosen ECG with no noise (at the top), 0.1 × *σ*, 0.5 × *σ* and 1 × *σ* (at the bottom).

**Figure 4.**
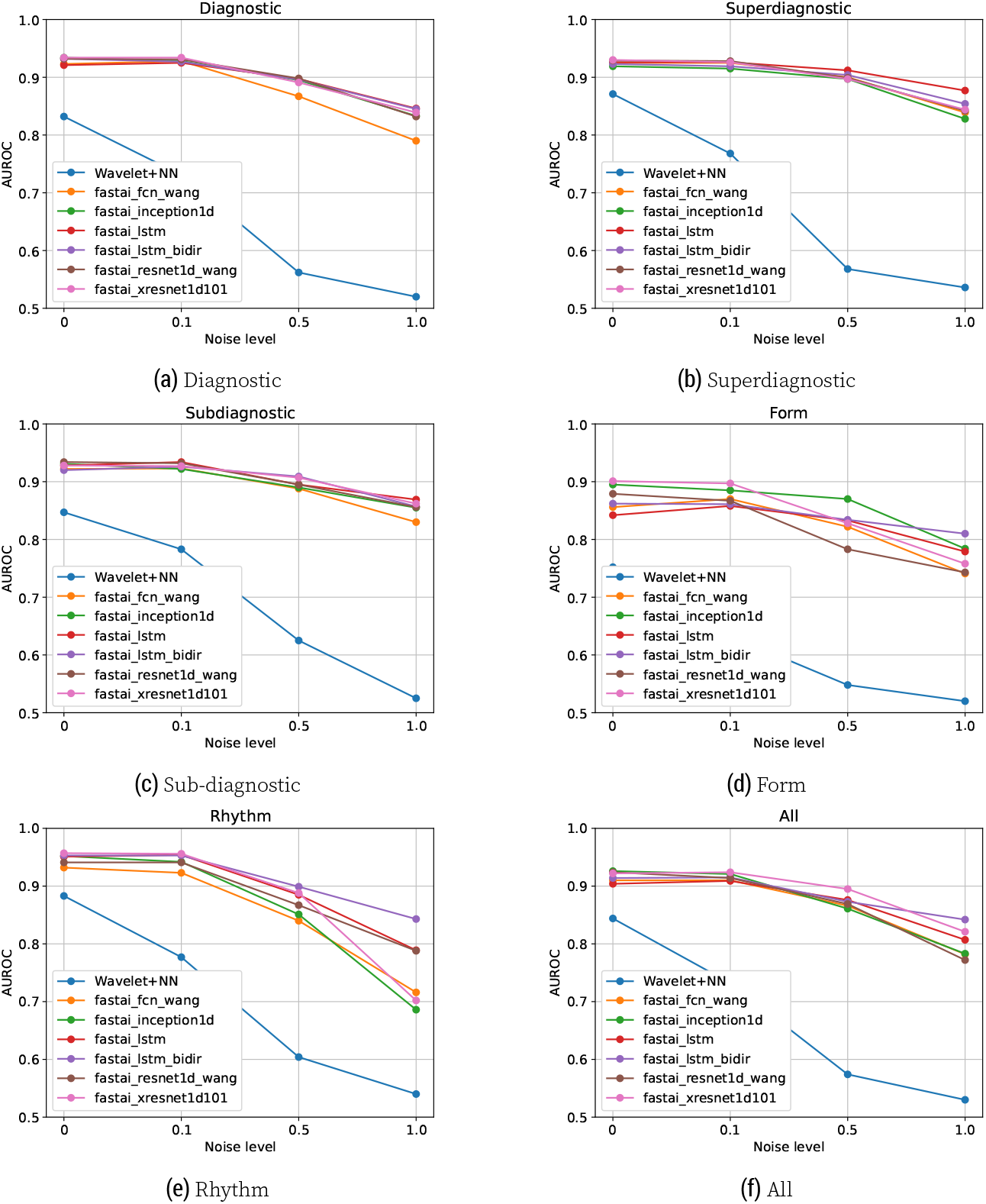
Performance, in terms of area under the receiver operating characteristic (AUROC), achieved by the seven different models on the six different benchmark tasks at different levels of noise added to the test data. The added stochastic noise where centered around zero with upper and lower boundaries given in terms of the standard deviation, *σ*, of all samples in the ECG test data multiplied by a coefficient *x*. In this study, we ran experiments with *x* = 0, 0.1, 0.5, and 1.

#### Proposing a new model

The optimal parameter combination of the Inception Time model, for each of the six benchmark tasks, is presented in Table 3. In addition to the different parameters, the table also presents the cross-validated score (3 folds) obtained on the subset of the training data that was used for hyperparameter optimization.

**Table 3.** Results from hyperparameter search Inception time.

| Parameter and score | All | diagnostic | subdiagnostic | superdiagnostic | form | rhythm |
| --- | --- | --- | --- | --- | --- | --- |
| Epoch | 15 | 25 | 15 | 25 | 25 | 25 |
| Batch size | 16 | 32 | 64 | 64 | 64 | 16 |
| Initial learning rate | 0.001 | 0.001 | 0.001 | 0.001 | 0.001 | 0.001 |
| Learning rate reduction | no | no | no | yes | no | no |
| Model depth | 9 | 6 | 6 | 12 | 6 | 9 |
| Loss function | *BCE | *BCE | **WBCE | *BCE | *BCE | **WBCE |
| Kernel size | (60,30,15) | (60,30,15) | (20,10,5) | (40,20,10) | (20,10,5) | (40,20,10) |
| AUROC score (mean 3 fold CV) | 0.887 | 0.870 | 0.878 | 0.895 | 0.800 | 0.915 |

\*BCE = Binary cross-entropy
\*\*BCE = Weighted binary cross-entropy

The final score on the test set, after training the models with the optimal configurations found in Table 3 on the whole training data set, are shown in Table 4

**Table 4.** Results achieved by the proposed Inception Time model on the six benchmark tasks with the optimal configurations found from grid search. The scores are given in terms of area under the receiver operating characteristic (AUROC) and the numbers in parenthesis represent the 95% confidential interval.

| Model | All | Diagnostic | Subdiagnostic | Superdiagnostic | Form | Rhythm |
| --- | --- | --- | --- | --- | --- | --- |
| Inception Time (all) | 0.926(08) |  |  |  |  |  |
| Inception Time (diagnostic) |  | 0.929(09) |  |  |  |  |
| Inception Time (subdiagnostic) |  |  | 0.927(08) |  |  |  |
| Inception Time (superdiagnostic) |  |  |  | 0.922(06) |  |  |
| Inception Time (form) |  |  |  |  | 0.840(11) |  |
| Inception Time (rhythm) |  |  |  |  |  | 0.923(32) |

## 5. Discussion

We successfully reproduced the most important results presented in Strodthoff et al. The results show that the mean of our repeated bootstrap experiments is within the 95% confidence interval presented in Strodthoff et al., with only 2 out of the 48 scores showing slight deviations. Despite these minor discrepancies, the core observations from the original paper remain valid. Additionally, we have confirmed the second claim put forth in the original paper. This claim entails the availability of a framework template that facilitates the straightforward implementation of new model architectures. Utilizing this template, we successfully created a new model and applied it to the benchmark tasks.

Interestingly the the end-to-end CNN performed well even with the additions of big noise portions to the ECG. Wavelet-based feature extraction combined with a dense neural network, on the other hand, had a steeper decline in performance when noise was added. A possible explanation is that some of the convolutional layers learn to suppress noise and effectively work as low/high-pass filters and thus perform better as a feature extractor than a static wavelet.

The Inception Time model proposed and implemented in this paper exhibits performance that closely matches that of the leading CNN models proposed by Strodthoff et al. across all six benchmark tasks. These results were somewhat contrary to our expectations since our proposed implementation of the Inception Time model had specialized configurations for each benchmark task. This shows that the models and the configurations proposed by Strodthoff et al. generally perform well across various ECG classification tasks. In future implementations and benchmark tests, one could try a larger search space in the grid search or potentially employ Bayesian hyperparameter tuning.

### 5.1 What was easy

When the code was successfully modified to be run in Google Colab it was easy to reproduce the results of Strodthoff et al. by simply running the reproduce_results.py script. It was also easy to add random noise to the ECGs in the test data to assess the model’s robustness to noise.

### 5.2 What was difficult

Implementing our proposed model using the templates in the GitHub repository took more time than we expected. We faced some errors when adding our proposed model to be tested on the benchmark tasks using the reproduce_results.py script. The error occurred because we oversaw some details in the configuration file. A more detailed explanation in the README file on how to add a custom model could mitigate future misunderstandings.

### 5.3 Communication with original authors

At the beginning of this replication study, we established communication with the authors of the paper. During the development and writing process, we got answers to all our questions and helped to submit our proposed model by performing a pull request to the original GitHub repository. The draft of this report was finally sent to the authors of the original paper to get their feedback before submission.

## Data Availability

All data produced are available online at:

https://doi.org/10.13026/6sec-a640

## Supplements

**Table 5.** Experiment number 1.

| Method | exp0_AUC | exp1_AUC | exp1.1_AUC | exp1.1.1_AUC | exp2_AUC | exp3_AUC |
| --- | --- | --- | --- | --- | --- | --- |
| fastai_inception1d | 0.927(06) | 0.928(12) | 0.925(14) | 0.917(06) | 0.892(12) | 0.960(10) |
| fastai_xresnet1d101 | 0.928(07) | 0.935(09) | 0.925(09) | 0.930(05) | 0.891(12) | 0.958(16) |
| fastai_lstm | 0.907(07) | 0.928(11) | 0.929(08) | 0.928(06) | 0.853(15) | 0.952(10) |
| fastai_lstm_bidir | 0.917(08) | 0.932(10) | 0.926(10) | 0.924(06) | 0.850(11) | 0.945(14) |
| Wavelet+NN | 0.834(13) | 0.831(17) | 0.846(23) | 0.871(08) | 0.763(18) | 0.878(22) |
| fastai_fcn_wang | 0.914(08) | 0.931(10) | 0.924(12) | 0.926(06) | 0.876(11) | 0.928(14) |
| fastai_resnet1d_wang | 0.918(06) | 0.933(10) | 0.930(09) | 0.929(06) | 0.881(18) | 0.946(10) |
| ensemble | 0.930(06) | 0.937(12) | 0.932(10) | 0.934(05) | 0.897(16) | 0.966(07) |
| naive | 0.500(00) | 0.500(00) | 0.500(00) | 0.500(00) | 0.500(00) | 0.500(00) |

**Table 6.**
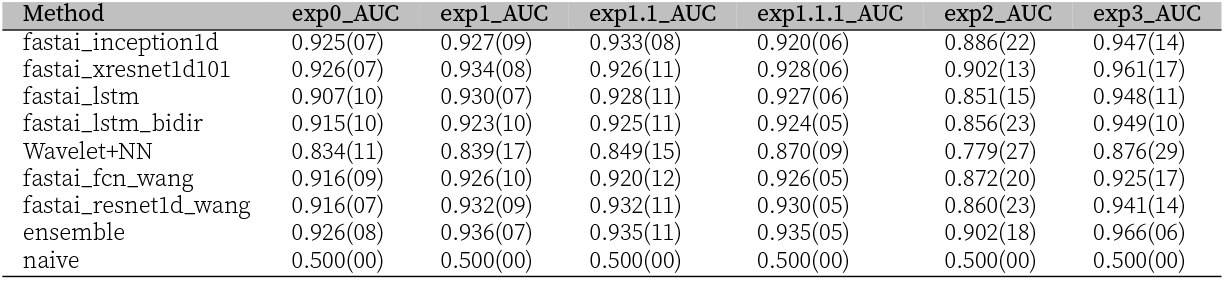
Experiment number 2.

**Table 7.**
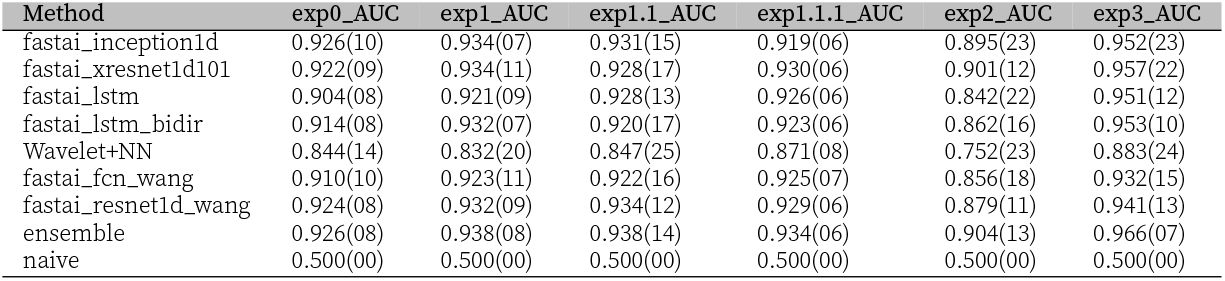
Experiment number 3.

## Footnotes

1 Source code Strodthoff et al. https://github.com/helme/ecg_ptbxl_benchmarking

2 The modified FastAI version is available here: https://github.com/Bsingstad/fastai

3 Our modified version of Strodthoff et al’s source code: https://github.com/Bsingstad/Strodthoff-2021

4 The PTB-XL dataset can be downloaded from here: https://physionet.org/content/ptb-xl/1.0.3/

## Notes

### Competing Interest Statement

The authors have declared no competing interest.

### Funding Statement

This study did not receive any funding

